# COVID-19 mortality and excess mortality among working-age Californians, by occupational sector: March 2020 through November 2021

**DOI:** 10.1101/2022.02.14.22270958

**Authors:** Yea-Hung Chen, Alicia R Riley, Kate A Duchowny, Hélène E Aschmann, Ruijia Chen, Mathew V Kiang, Alyssa Mooney, Andrew C Stokes, M Maria Glymour, Kirsten Bibbins-Domingo

## Abstract

**Background:** During the first year of the pandemic, essential workers faced higher rates of SARS-CoV-2 infection and COVID-19 mortality than non-essential workers. It is unknown whether disparities in pandemic-related mortality across occupational sectors have continued to occur, amidst SARS-CoV-2 variants and vaccine availability.

**Methods:** We obtained data on all deaths occurring in the state of California from 2016 through 2021. We restricted our analysis to California residents who were working age (18–65 years at time of death) and died of natural causes. Occupational sector was classified into 9 essential sectors; non-essential; or not in the labor market. We calculated the number of COVID-19 deaths in total and per capita that occurred in each occupational sector. Separately, using autoregressive integrated moving average models, we estimated total, per-capita, and relative excess natural-cause mortality by week between March 1, 2020, and November 30, 2021, stratifying by occupational sector. We additionally stratified analyses of occupational risk into regions with high versus low vaccine uptake, categorizing high-uptake regions as counties where at least 50% of the population completed a vaccination series by August 1, 2021.

**Findings:** From March 2020 through November 2021, essential work was associated with higher COVID-19 and excess mortality compared with non-essential work, with the highest per-capita COVID-19 mortality in agriculture (131.8 per 100,000), transportation/logistics (107.1), manufacturing (103.3), and facilities (101.1). Essential workers continued to face higher COVID-19 and excess mortality during the period of widely available vaccines (March through November 2021). Between July and November 2021, emergency workers experienced higher per-capita COVID-19 mortality (113.7) than workers from any other sector. Essential workers faced the highest COVID-19 mortality in counties with low vaccination rates, a difference that was more pronounced during the period of the Delta surge in Summer 2021.

**Interpretation:** Essential workers have continued to bear the brunt of high COVID-19 and excess mortality throughout the pandemic, particularly in the agriculture, emergency, manufacturing, facilities, and transportation/logistics sectors. This high death toll has continued during periods of vaccine availability and the delta surge. In an ongoing pandemic without widespread vaccine coverage and anticipated threats of new variants, the US must actively adopt policies to more adequately protect essential workers.

## Introduction

Individuals working in essential occupations (ie, in sectors deemed essential to local or regional functions and thus exempt from public health stay-at-home orders or other restrictions to in-person work) have faced higher risk of SARS-CoV-2 infection and COVID-19 mortality during the pandemic.^1–4^ An analysis of data from the UK Biobank project, for example, found higher risk of severe COVID-19 disease among essential workers than among non-essential workers.^2^ Another study using data from the American Community Survey linked to mortality records from the Social Security Administration found that people without work-from-home options had larger increases in mortality during 2020 than people working in occupations that had the option to work from home.^5^ It is unknown whether such disparities have continued to occur amidst SARS-CoV-2 variants and vaccine availability.

We previously reported on COVID-19 and excess mortality by occupation in California from the start of the pandemic (March 2020) through November 2020.^1^ We found that essential work was associated with higher COVID-19 and excess mortality during this period compared with non-essential work and that four essential sectors—food and agriculture, manufacturing, transportation and logistics, and facilities— experienced particularly high excess mortality. The present study adds three major updates. One, we extend the time window of interest through November 2021, through an era that includes SARS-CoV-2 variants and vaccine availability. Two, we further disaggregate data for two essential sectors sectors we have previously combined: health/emergency and food/agriculture. We note here that different policies and behaviors between sectors—such as between health workers and emergency workers (first responders)—may have translatated to differences in risk. Three, we report on differences in COVID-19 mortality between regions of low or high vaccination.

## Methods

We obtained data from the California Department of Public Health on deaths occurring in the state from 2016 through 2021. We restricted our analyses to California residents who were 18–65 years (inclusive of endpoints) at the time of death and died of natural causes. We restricted to natural-cause deaths so that our estimates of excess mortality would more plausibly identify unrecognized or unrecorded COVID-19 deaths.

We identified recorded COVID-19 deaths by searching through all 20 cause-of-death variables available to us. Hereafter, for brevity, we may use “COVID-19 deaths” in place of “recorded COVID-19 deaths” and “excess deaths” in place of “excess natural-cause deaths.”

Data on occupation are recorded on death certificates via free-text responses. For example, a family member or friend might indicate that a decedent was a “cook” during most of the decedent’s life. We converted these free-text data to US Census codes using the National Institute for Occupational Safety and Health’s Industry & Occupation Computerized Coding System. A team of 3 researchers then categorized each unique code into one of 11 occupational sectors: agriculture, emergency services, facilities, government/community, health, manufacturing, restaurant, retail, transportation/logistics, not essential, and unemployed/missing (this category includes homemakers, retirees, and students). Our choice of sectors was guided by the 13 sectors identified by California officials as comprising the state’s essential workforce.^6^

Our time period of interest is March 1, 2020 through November 30, 2021. In time-stratified analysis, we divided the time window into four phases: March through November 2020 (Phase 1), December 2020 through February 2021 (Phase 2), March 2021 through June 2021 (Phase 3), and July through November 2021 (Phase 4). Vaccines became available to health workers in California near the beginning of the second phase,^7^ while emergency workers and other essential workers became eligible near the beginning of the third phase.^8^ The first and fourth phases correspond to surges in COVID-19 cases and deaths.

In a secondary analysis, we stratified by California counties with low or high vaccine uptake among working-age individuals, using data from the Centers for Disease Control and Prevention.^9^ We defined counties with low vaccination as counties with 50% of the population fully vaccinated by August 1, 2021. While this definition was somewhat arbitrary, we note here that differentiation between the high and low counties—as explored via time series of the vaccine uptake in each county (not shown)—was consistent over time.

For each analytic group of interest (usually the entire state or a specific occupational sector), we repeated the following procedure. We calculated the number of COVID-19 deaths occurring in total and per capita in each week, over the entire time window, and in each phase. We obtained subgroup-specific population estimates, for the per-capita numbers, from the 2019 American Community Survey. Separately, we fit dynamic harmonic regression models with autoregressive integrated moving average errors for the number of weekly deaths,^10^ using deaths occurring among the group between January 3, 2016 and February 29, 2020. Using the final model, we forecast the number of deaths for each unit of time, along with corresponding 95% prediction intervals (PI). To obtain the total number of excess deaths for the entire time window and during each phase, we subtracted the total number of expected (forecast) deaths from the total number of observed deaths. We obtained a 95% PI for the total by simulating the model 10,000 times, selecting the 97.5% and 2.5% quantiles, and subtracting from the total number of observed deaths.

In addition to the estimated number of excess deaths, we calculated and report per-capita excess deaths: the observed number of deaths minus the expected number of deaths, divided by the estimated population size. We also calculated and report the observed number of deaths divided by the expected number of deaths. These ratios represent relative excess mortality. For example, a ratio of 1.5 would indicate that there were 50% more deaths observed during the pandemic than we would have expected had the pandemic not occurred. For both measures, the comparison is between the pandemic and non-occurrence of the pandemic. The reference group is non-occurrence of the pandemic.

We focus our presentation of our findings on COVID-19 mortality, and view the estimates of excess mortality as sensitivity checks of the COVID-19 numbers, particularly given discrepancies between COVID-19 and estimated excess mortality early in the pandemic (which we believe to be primarily due to unrecognized or unreported COVID-19 deaths).

Our reported per-capita numbers are annualized numbers, obtained by dividing the per-capita measure by the number of the weeks and multiplying by 52.

Because differences between occupational sectors could be impacted by differences in age and sex, we also performed age- and sex-stratified sensitivity analyses.

Our use of the death data was approved by the State of California Committee for the Protection of Human Subjects.

We conducted all analyses in R.

## Results

Between March 2020 and November 2021, there were 24,799 COVID-19 deaths reported among working-age Californians and an estimated 28,751 (95% PI: 27,853 to 29,653) excess deaths (Table 1). Across occupational groups, workers in the agriculture, transportation/logistics, manufacturing workers, and facilities sectors experienced the highest per-capita COVID-19 mortality, per-capita excess mortality, and relative excess mortality. For example, among agriculture workers, there were 131.8 reported COVID-19 deaths per 100,000 and an estimated 159.3 (95% PI: 149.6 to 169.1) excess deaths per 100,000; the relative excess mortality among the group was 1.61 (95% PI: 1.55 to 1.67). Workers in non-essential sectors experienced the lowest per-capita COVID-19 mortality (27.5), per-capita excess mortality (27.1; 95% PI: 16.4 to 37.5), and relative excess mortality (1.17; 95% PI: 1.10 to 1.26).

**Table 1:**
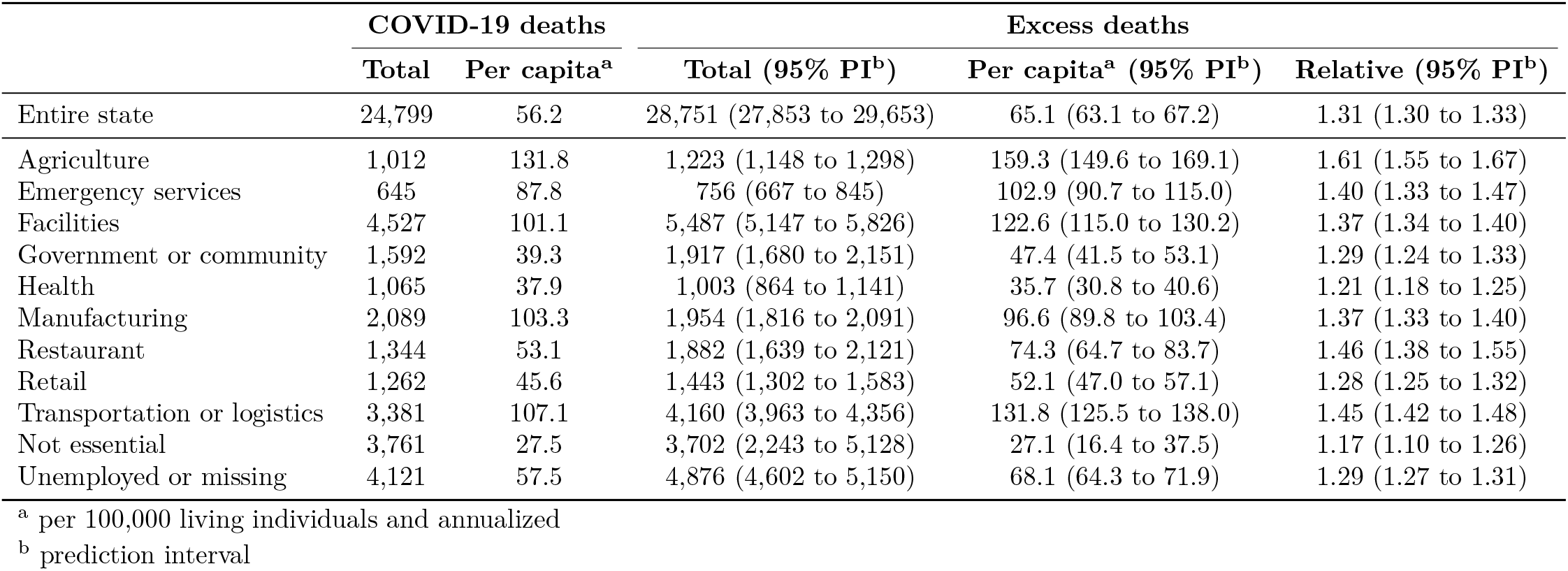
Excess natural-cause mortality among Californians 18–65 years of age, by occupational sector, March 2020 through November 2021.

Per-capita COVID-19 mortality varied over time, peaking during Winter 2020–2021 and Summer 2021 (Figure 1 and Table 2). During the Winter 2020–2021 surge, per-capita COVID-19 mortality was highest among workers in the agriculture (373.8), manufacturing (367.8), transportation/logistics (350.8), and facilities (330.9) sectors, and lowest among workers in non-essential sectors (85.1). During the Summer 2021 surge, per-capita COVID-19 mortality was highest among workers in the emergency (113.7), agriculture (111.5), transportation/logistics (105.3), and facilities (96.5) sectors, and lowest among workers in non-essential sectors (33.6).

**Table 2:** Per-capita^a^ COVID-19 mortality (and rank for the five riskiest sectors) among Californians 18–65 years of age, by occupational sector and phase, March 2020 through November 2021.

|  | Mar to Nov 2020 | Dec 2020 to Feb 2021 | Mar 2021 to Jun 2021 | Jul 2021 to Nov 2021 |
| --- | --- | --- | --- | --- |
| Entire state | 29.9 | 182.9 | 20.1 | 55.8 |
| Agriculture | 103.6 (1) | 373.8 (1) | 37.6 (2) | 111.5 (2) |
| Emergency services | 40.0 (5) | 269.7 (5) | 24.8 (5) | 113.7 (1) |
| Facilities | 55.6 (4) | 330.9 (4) | 36.0 (3) | 96.5 (4) |
| Government or community | 18.1 | 130.9 | 15.5 | 41.3 |
| Health | 21.9 | 119.9 | 12.2 | 37.9 |
| Manufacturing | 61.4 (2) | 367.8 (2) | 29.6 (4) | 78.1 (5) |
| Restaurant | 30.2 | 186.5 | 20.1 | 40.2 |
| Retail | 21.6 | 143.8 | 16.4 | 52.6 |
| Transportation or logistics | 55.9 (3) | 350.8 (3) | 40.5 (1) | 105.3 (3) |
| Not essential | 12.2 | 85.1 | 10.9 | 33.6 |
| Unemployed or missing | 31.3 | 189.2 | 20.6 | 54.8 |

<sup>a</sup> per 100,000 living individuals and annualized

**Figure 1:**
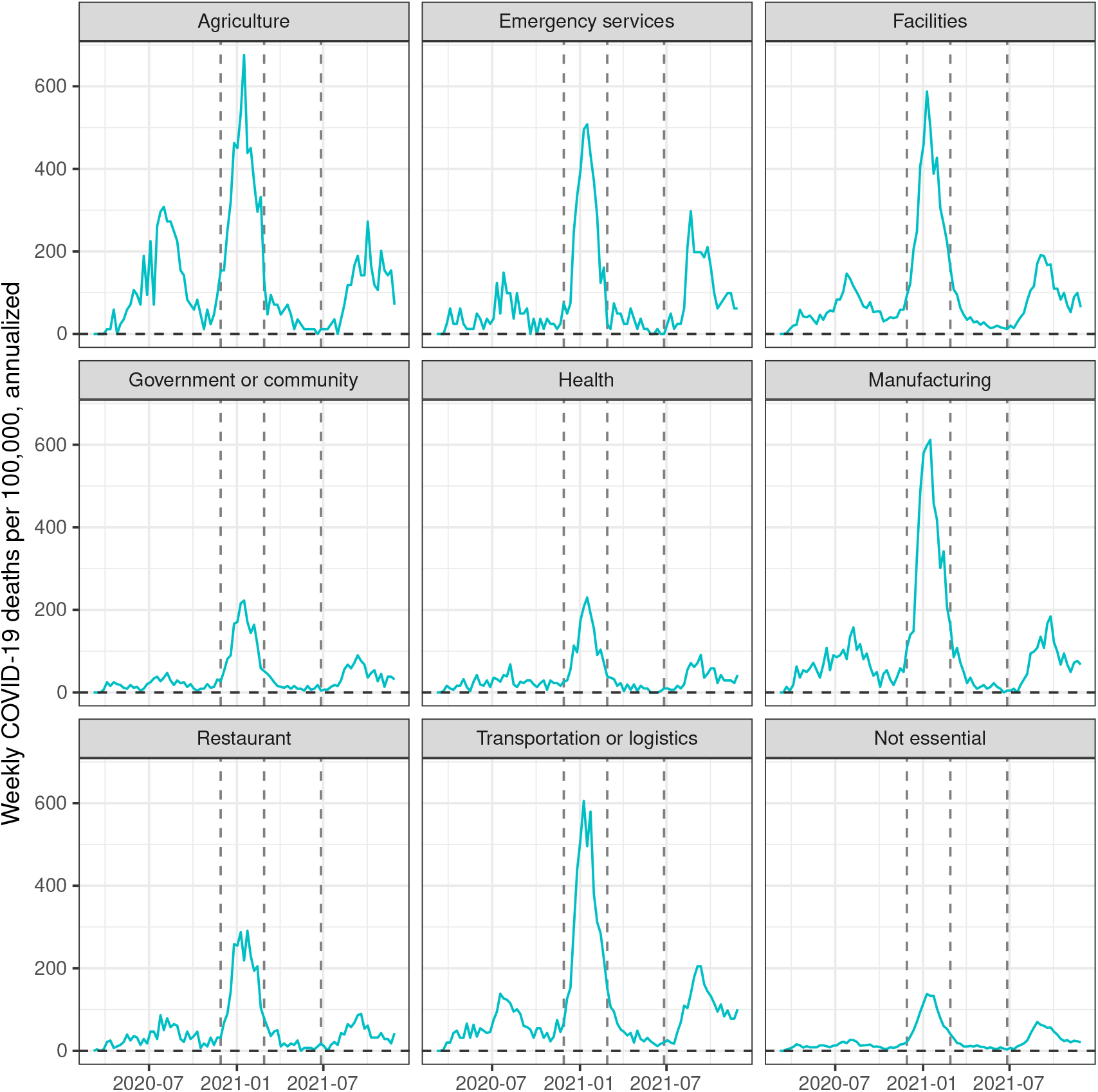
Per-capita COVID-19 mortality among Californians 18–65 years of age, by occupational sector, March 2020 through November 2021.

In the post-Winter phase, from March through June 2021, per-capita COVID-19 mortality was comparable between regions that subsequently had low and high vaccine uptake, regardless of occupational sector (Figure 2). However, COVID-19 mortality in low and high vaccination regions diverged during the Summer 2021 surge. Among essential workers other than health workers, the difference in annualized per-capita COVID-19 mortality between high and low vaccination regions was 9.6 per 100,000 individuals prior to the Summer 2021 surge and 78.8 per 100,000 individuals during the Summer 2021 surge.

**Figure 2:**
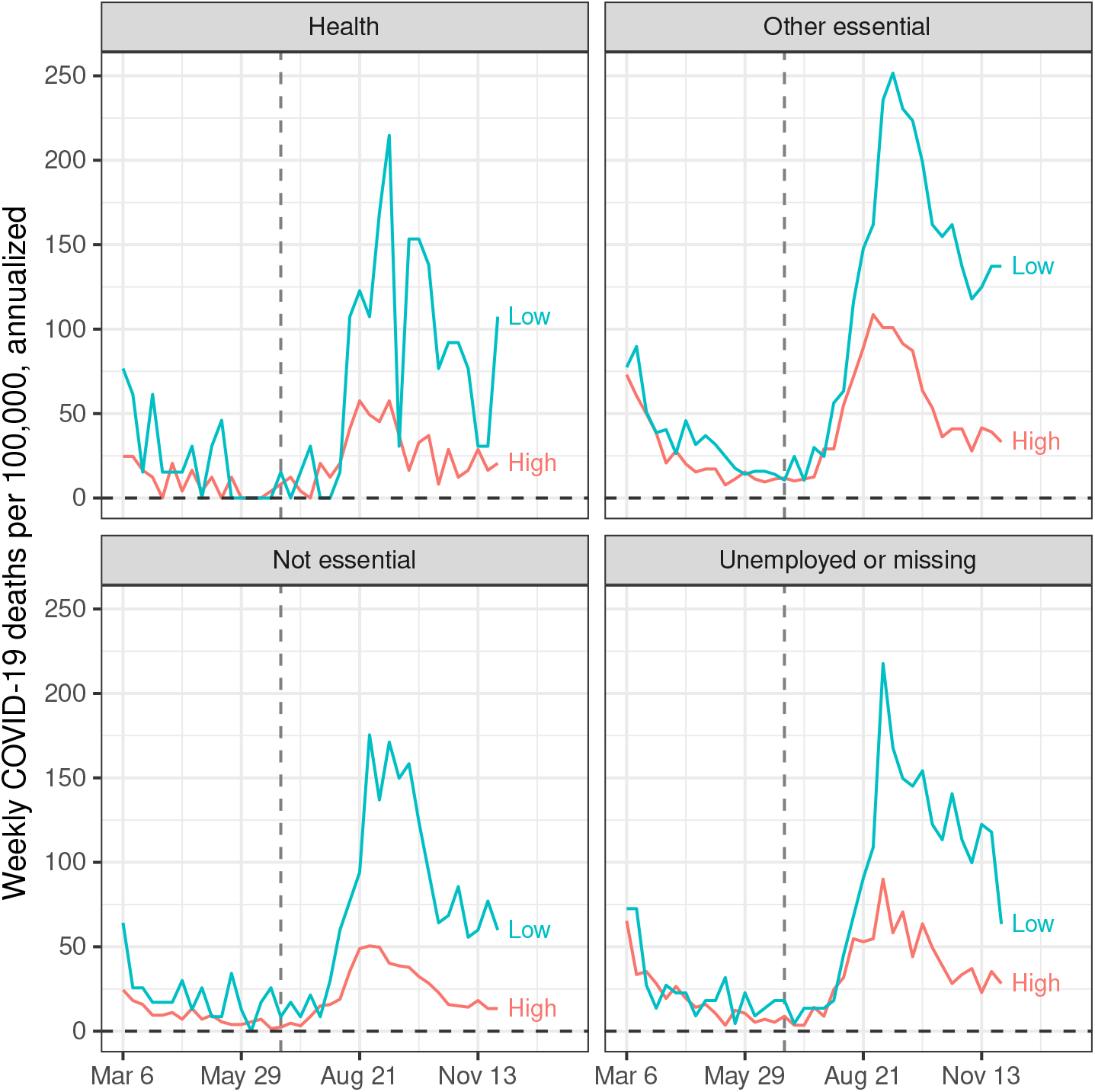
Per-capita COVID-19 mortality among Californians 18–65 years of age, by occupational sector and regions of low or high vaccine uptake, March through November 2021.

In sensitivity analyses, we further stratified our analysis of COVID-19 deaths by sex (male/female) and by age group (18–35, 36–55, and 56–65), finding no meaningful changes to our findings of higher per-capita COVID-19 mortality among essential workers. For example, among individuals 18–35 years of age, annualized per-capita COVID-19 mortality was highest among workers in the emergency (15.4), transportation/logistics (13.0), and manufacturing (11.4) sectors, and lowest among non-essential workers (3.6). Similarly, among individuals 56–65 years of age, per-capita COVID-19 mortality was highest among workers in the agriculture (468.1), transportation/logistics (330.4), and manufacturing (327.3) sectors, and lowest among non-essential workers (85.2). Among females, per-capita COVID-19 mortality was highest among workers in the agriculture (60.4), manufacturing (54.2), and emergency (48.1) sectors, and lowest among non-essential workers (21.3). Among males, per-capita COVID-19 mortality was highest among workers in the agriculture (168.4), manufacturing (124.7), and transportation/logistics (123.4) sectors, and lowest among non-essential workers (34.1).

## Discussion

This comprehensive analysis of COVID-19 and excess mortality by occupational sector in California from March 2020 to November 2021 yields three important sets of findings. First, essential work remains associated with far higher COVID-19 and excess mortality compared with non-essential work, with the highest mortality among workers in the sectors of agriculture, emergency services, facilities, manufacturing, and transportation/logistics. Through November 2021, the risk of death was highest among agricultural workers, with mortality 61% higher than expected based on prior periods. Second, essential workers continued to face high COVID-19 and excess mortality during the period of widely available vaccines (March–November 2021). Notably, during the Summer 2021 surge, workers in emergency services faced the highest per-capita COVID-19 mortality of all the essential sectors. Finally, both occupational sector and level of county vaccine coverage are associated with COVID-19 mortality. Among essential workers, per-capita COVID-19 mortality was higher among residents in high-vaccine regions than among residents in low-vaccine regions, particularly during the Summer 2021 surge.

In this study, per-capita COVID-19 mortality was 2.2 times higher among emergency workers than among health workers during the Winter 2020–2021 surge; this ratio grew to 3.0 during the Summer 2021 surge. We hypothesize that this widened disparity between emergency and health workers during Summer 2021 reflects, in part, low levels of vaccine uptake among emergency workers and high levels of vaccine uptake among health workers. In Los Angeles, the state’s most populous city, just over 50% of police officers or firefighters were at least partially vaccinated by mid June 2021.^11^ In comparison, roughly 77% of employees across 350 state hospitals were at least partially vaccinated by mid July 2021; vaccine mandates for the group are believed to be a contributing factor to the relatively high uptake.^12^ Our study suggests that vaccination may be particularly crucial during surge periods. During the Summer 2021 surge, the peak per-capita COVID-19 mortality among non-health essential workers was 251.6 in low-vaccination regions and 108.6 in high-vaccination regions, corresponding to a relative disparity of 2.3.

Protecting workers whose jobs are essential to critical functions and infrastructure should be a priority in the pandemic response. Yet, deaths among essential workers remain high. While vaccine prioritization for essential workers has likely been highly effective in prolonging lives (as suggested, for example, in our analysis stratifying by regions of low of high vaccination), it is clear that uptake has been insufficient to reduce disparities. Efforts to increase vaccine uptake among workforces via mandates have been perceived as effective in several cases,^13,14^ but have faced legal challenges. Notably, the US Supreme Court has ruled against the Occupational Safety and Health Administration’s emergency temporary standard requiring vaccination or testing at businesses with at least 100 employees.^15,16^ And, in the City of Los Angeles, groups of police officers and firefighters have filed lawsuits in opposition to the city’s vaccine mandate.^17,18^ These legal decisions and battles represent meaningful challenges to reducing COVID-19 mortality among essential workers in the US.

In the absence of a national mandate, the nation will have to lean on other strategies. These may include local or private vaccine mandates, such as the vaccine mandates at Tyson Foods and United Airlines.^13,19^ Community-based or employer-sponsored^20^ vaccination efforts can address structural barriers (such as limited access to transportation) and misinformation.^21,22^ Policies can and should address the unique challenges and risks that low-income individuals face during the pandemic,^23^ including job security, financial burdens of healthcare, and disruptions of schooling. Paid sick leave, for example, can ensure that essential workers do not have to choose between financial benefits and health risks.^24^ Finally, protections in workplace settings remain crucial. Given that SARS-CoV-2 can be transmitted via aerosols,^25,26^ we urge for free provision of masks—preferably N95 masks or similarly effective masks^27,28^—to essential workers and improved ventilation in workplace settings.^29^

We acknowledge challenges related to measurement of occupation on death certificates, and questions relating to whether risks are necessarily related to workplace exposures. These are addressed in our prior manuscript at length.^1^

More than one year after we first reported disparities in COVID-19 mortality across occupational sectors,^1^ and even after widespread availability of vaccination, these disparities continue to occur. The patterns are clear: though there have been a large number of COVID-19 deaths among non-essential workers, essential workers continue to bear the brunt of the COVID-19 pandemic. However, not all essential workers face the same elevated risk. Contrary to popular messaging, health sector workers faced lower risk of COVID-19 and excess mortality than workers in several non-health sectors (agriculture, emergency, facilities, manufacturing, and transportation/logistics) both prior to and after vaccine availability. These findings challenge notions about the basis of occupational risk during the pandemic, suggesting that increased occupational risk of COVID-19 death may have more to do with workplace safety, worker protections, and worker power, than mere proximity to COVID-19. Further, even in counties with high vaccination, essential sectors experienced elevated risks of COVID-19 mortality and excess mortality, suggesting that vaccine uptake alone has been insufficient to erase previously documented disparities in COVID-19 death. We urge for decisive and collaborative action, using a diverse toolkit, to reduce and eliminate disparities across occupational groups.

## Data Availability

All data produced are available via application to the State of California

https://www.cdph.ca.gov/Programs/CHSI/Pages/Program-Landing1.aspx

